# Peer Education Intervention Reduced Sexually Transmitted Infections Among Male Tajik Labor Migrants Who Inject Drugs: Results of a Cluster-randomized Controlled Trial

**DOI:** 10.1101/2024.08.15.24312070

**Authors:** Mary Ellen Mackesy-Amiti, Judith A. Levy, Casey M. Luc, Jonbek Jonbekov

## Abstract

**Background:** Male Tajik labour migrants who inject drugs in Russia are at high risk for HIV and other sexually transmitted infections. The “Migrants’ Approached Self-Learning Intervention in HIV/AIDS for Tajiks” (MASLIHAT) trained Tajik labour migrants who inject drugs in Moscow as peer educators (PEs) in delivering HIV prevention information and promoting risk-reduction norms and practices within their diaspora social networks while reducing their own HIV risk. Our earlier analysis of a cluster-randomized controlled trial testing the intervention’s effects found that MASLIHAT reduced condomless sex, condomless sex with female sex workers, and sex with multiple sexual partners. This analysis draws on data from this parent study to investigate if these observed changes in safer sex translated into fewer reported STIs over 12-months.

**Methods:** Male Tajik migrant workers in Moscow who inject drugs (n=140) were recruited from construction worksites, local bazaars, and diaspora organizations serving labor migrants. Participants were assigned as PEs to either MASLIHAT or a comparison health education intervention. Each PE recruited two migrants who inject drugs from their social networks with whom to share what they learned during the 5 educational sessions of the arm to which they were assigned. All participants completed questionnaires at baseline and 3-month intervals for one year to assess their HIV/STI risk behaviour. Mixed effects robust Poisson regression analyses tested for possible differences between assignment conditions in self-reported STIs during 12 months of follow-up and the contribution of sexual risk behaviours to STI acquisition. We then tested the mediating effects of sexual behaviours during the first six months following the intervention on STIs reported at the 9 and 12-month follow-up.

**Results:** Participants in the MASLIHAT intervention were significantly less likely to report an STI during follow-up (IRR=0.27). Condomless sex with a casual or commercial partner was significantly associated with STI acquisition (IRR=2.30). Causal mediation analysis indicated that the intervention’s effect on reported STI was partially mediated by reductions among MASLIHAT participants in condomless sex with a casual or commercial partner.

**Conclusions:** The MASLIHAT peer-education intervention reduced reported STIs among Tajik labour migrants partly through reduced condomless sex with casual and commercial partners.

**Clinical trial registration:** ClinicalTrials.gov, 2021-04-16, NCT04853394.

## Background

Sexually transmitted Infectious diseases (STIs) have become a rapidly growing public health problem worldwide. The World Health Organization (WHO) estimates that globally more than a million curable new STD infections occur each day (1). When left untreated, STIs can lead to serious long-term health outcomes including genital and other cancers, pelvic inflammatory disease, pregnancy complications such as pre-term delivery and low neonatal birth weight, infertility, and neurologic manifestations.(2) STIs also facilitate HIV acquisition by causing a break in the skin that offers easier access for the virus to enter the body (3), and they also facilitate forward transmission by increasing viral concentration in the genital tract (4). STIs are frequently used in research and clinical practice as a marker for HIV risk, (6) as they share many of the same sexual risk behaviors. STIs themselves however represent a critical problem that deserves greater attention than they currently receive. (4)

This analysis addresses the underrepresentation of STIs in the scientific literature by investigating whether a successful HIV risk-reduction intervention for Tajik labor migrants in Moscow who inject drugs reduced their incidence of STIs over a one-year period. Almost one million Tajik citizens work as temporary labor migrants outside the country each year with Russia the major host destination, a terminus with an exceedingly high HIV prevalence (7) despite a recent somewhat downward trend in incidence.(8) Some large but unknown numbers of Tajik migrants inject drugs while in Russia, either as newly introduced or long-time users.(9) As a temporary labor force, Tajik migrants perform many of Moscow’s most dangerous jobs in construction and the transportation of goods, sales, and food service in bazaars and markets. Typically living in crowded, substandard housing at a bare subsistence level, they are highly vulnerable to feelings of loneliness, depression, and general anxiety plus chronic and infectious diseases.(10) Meanwhile, access to medical treatment for migrants in Russia is limited.(11) As true among labor migrants globally (12, 13), these factors contribute to many of these migrant males engaging in risky sex without a condom, with multiple partners, and with female sex workers (FSWs) in Moscow.(9, 12, 14) FSW in Moscow have prevalence of HIV exceeding 3% as well as other STIs.(15)

To address the need for both HIV and STI prevention for this population we developed and tested the Migrants’ Approached Self-Learning Intervention in HIV/AIDS for Tajiks (MASLIHAT), a peer-network educational intervention for reducing risky sex, drug, and alcohol, behavior among Tajik migrants who inject drugs. Peer-education models have demonstrated a long and successful history of reducing HIV and related diseases among PWID beginning as far back as the late 1980s (16, 17) through to recent years.(18, 19) In a cluster-randomized parallel group controlled trial, the MASLIHAT peer intervention demonstrated significant effects in reducing risky sex and injection drug behavior among both peer educator participants (PEs) and their network members (NMs). (20, 21)

For our current analysis, we turn our attention to answering a critical intervention outcome question, “Did MASLIHAT’s observed reduction in sexual risk behavior lower the incidence of STIs among this population of male Tajik migrants in Moscow?” To this end, we test the hypotheses that (1) MASLIHAT participants would report fewer STIs during one-year of follow-up compared to TANSIHAT participants and (2) the effect of the intervention on reported STIs is mediated by changes in sexual risk behavior.

## Methods

Study procedures were reviewed and approved by the Institutional Review Boards of the University of Illinois Chicago (protocol 2020-0795), PRISMA Research Center in Tajikistan, and the Moscow Nongovernment Organization “Bridge to the future.” All participants provided written informed consent. The MASLIHAT trial was registered with ClinicalTrials.gov, NCT04853394.

### Recruitment and Site Assignment

From October 2021 to April 2022, 140 male Tajik migrant workers who inject drugs were recruited in Moscow through referral from two Tajik diaspora organizations and a call for study volunteers disseminated by Tajik managers and work brigade leaders at 10 bazaars or construction sites that employ large numbers of Tajik migrants. (20). Prospective participants had to be a male Tajik migrant aged 18 or older, a current or former person who injects drugs (PWID), intending to reside in Moscow for the next 12 months to participate in their assigned intervention and follow-up data collection, and willing to recruit two male PWID to participate as IDU network members (NMs) for baseline and follow-up surveys. Network members (n=280) had to meet the same eligibility criteria as directly recruited participants but also: 1) have injected drugs at least once in the last 30 days; and 2) be someone whom their recruiter sees at least once a week to facilitate information diffusion.

Recruitment sites were paired according to socio-demographic characteristics, and each site randomly assigned using a random number generator to deliver one of the two intervention arms. Only the study’s PI and project director knew which sites were assigned to which intervention, and sites were sufficiently distant geographically to avoid intervention cross-contamination. Directly recruited participants were assigned post-recruitment as PEs to the intervention arm in their recruitment area. The study’s overall sample consists of 140 PEs and 280 network members for a total of 420 participants. Participants received the equivalent of $20.00 in Russian Rubles for their time and transportation costs in participating in each intervention session (PEs only) and for completing surveys at baseline and follow-up (both PEs and NMs). A CONSORT diagram is provided in an additional file [see Additional File 1].

### Intervention sessions

MASLIHAT is a small-group, interactive intervention that uses peer networks to reduce drug, alcohol, and sexual risk behaviors among temporary migrant workers who inject drugs. Labor migrants in the host country who currently or have previously injected drugs are trained as PEs to promote positive HIV risk-reduction norms and behavioral change through role modeling and by sharing what they learned during MASLIHAT educational sessions with their at-risk network members. The intervention includes five HIV knowledge and skill-building sessions that involve goal setting, role playing, demonstrations, homework, and group discussions. These sessions teach participants techniques for personal HIV and STI risk reduction along with the communication and outreach skills needed to encourage others at risk also to adopt them. The TANSIHAT program echoes MASLIHAT’s 5 sessions in style and time commitment but focuses instead on healthy living without any content related to HIV or STI sexual risk behavior. The intervention sessions per arm were delivered by different facilitators in groups of 4−7 at the PRISMA Research Center in Moscow. Sessions were scheduled weekly and lasted 2 hours.

### Baseline and follow-up surveys

Baseline surveys with PEs and NMs were conducted at the PRISMA office in Moscow or a private location of the participant’s choosing. Following the survey, participants in both conditions were referred to the Moscow HIV Prevention Center to be tested for HIV and hepatitis C virus. Anonymized test results were reported to study staff with only a group number to identify the recruitment site. Follow-up surveys were conducted with PEs and NMs in both arms at 3-month intervals over 12 months.

### Measures

The structured baseline and follow-up questionnaires collected information on sociodemographic characteristics, drug-related practices, and sexual behavior. As our current analysis focuses solely on sexual behavior and STI acquisition and not drug use, only these measures are described below.

#### Sexually transmitted infections

At each follow-up, participants were asked if they had been diagnosed in the past three months with gonorrhea, syphilis, chlamydia, or any other STIs. Responses were coded as yes or no to indicate a STI diagnosed in the past three months. While testing periodically for its occurrence, HIV is not included in measuring STI prevalence as our intent was to examine STIs independent of HIV – a much neglected issue in the scientific literature.(4)

#### Sexual risk behavior

Participants were asked as to how many women in Moscow with whom they had sexual intercourse in the past 30 days, and how many of these partners were a regular main partner, FSW, or casual sexual partners not engaged in selling sex. Responses were used to create binary measures of multiple female partners and any FSW partner in the past 30 days. Condom use was assessed by asking participants, “How often did you use a condom when having sexual intercourse?” for each of three partner categories. Response categories were “never,” “sometimes,” “often,” or “always.” Responses were coded to create binary measures of any condomless sex (CS), condomless sex with FSWs (CS/FSW), and condomless sex with a casual or commercial (i.e. FSW) partner (CS/CC) in the past 3 months.

### Analysis

We conducted mixed effects logistic and modified Poisson regression analyses predicting sexual risk behaviors, with random intercepts for participant, network cluster, and site to obtain estimates for the interaction effect of treatment arm and time. We selected logistic models for outcomes with low prevalence during follow-up. We first estimated null models to test for clustering effects. If variance at the site level was not significant, site level clustering was omitted from the model. Time was included as four dummy variables for 3, 6, 9, and 12-month follow-up vs. baseline. Marital status (married vs. not currently married) and income were included as covariates, as they varied significantly across arms.

Using follow-up data, we conducted mixed effects logistic regression models predicting any STI, with random intercepts for participant, network cluster, and site to obtain estimates for the effects of treatment arm and sexual risk behaviors. Time was initially included as three dummy variables for 6, 9, and 12-month vs. 3-month follow-up, and we tested the interaction of condition and time. Non-significant effects (p>.10) were not included in subsequent models. In separate models, we tested sex with FSWs, CS, CS/FSW, CS/CC and sex with multiple partners to assess the effect of treatment arm when adjusted for these behaviors. Analyses were conducted using Stata (version 18).

We estimated a causal mediation model (Stata *medeff*) to compute the indirect effects of treatment arm on STI reported at 9- and 12-month follow-up via sexual risk behaviors reported at 3- and 6-month follow-up (Figure 1). Robust standard errors were computed to adjust for network clustering. Only sexual risk behaviors that were significantly affected by the intervention, and significantly associated with STI at later follow-up were considered as potential mediators. We included the effect of baseline behavior and tested the interaction between baseline behavior and treatment arm. We also included covariates that were found to be significantly associated with the mediator or outcome.

**Figure 1.**
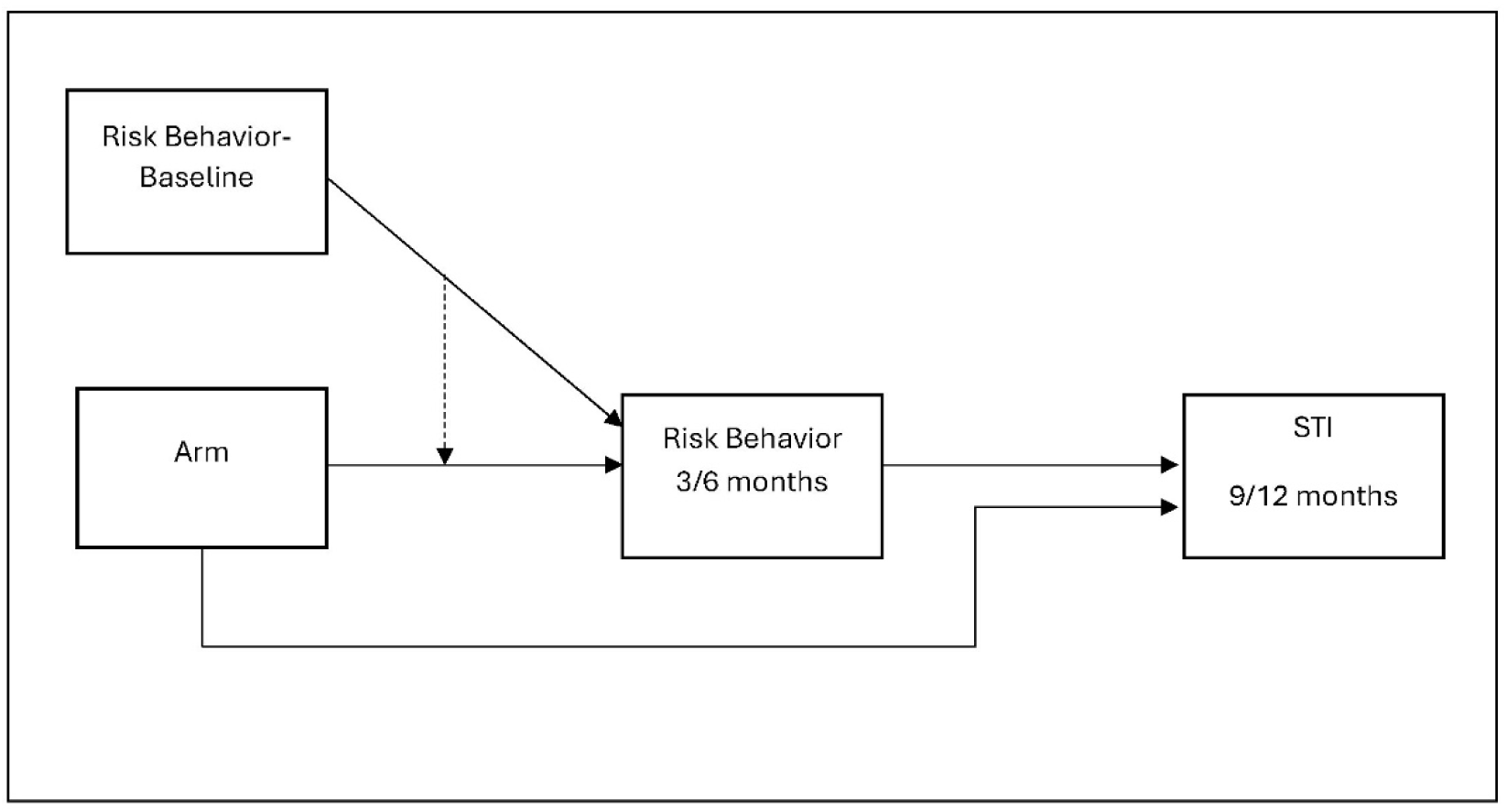
Mediation model of treatment arm effect on STI at 9 or 12-month follow-up by sexual risk behavior at 3 or 6-month follow-up.

## Results

Table 1 reports the demographic characteristics of the sample. All five major regions of Tajikistan (the capital city of Dushanbe, Region of Republican Subordination, Khatlon, Sugd, and Gorno-Badakshan Autonomous Oblast) are represented in the sample. More participants in the control condition were married (Chi2=5.62, p=0.014), and their income was lower (t=2.42, p=0.016) compared to the MASLIHAT condition. HIV prevalence was 6.8%, among participants who agreed to testing (n=413). When asked at baseline if they had ever been diagnosed with an STI, 41% of the total sample reported yes, 49% reported no, and 10% declined to answer. There was no significant difference in lifetime STI rates between treatment arms at baseline (IRR=0.98, p=0.92). Lifetime STI was significantly associated with employment sector (chi2[3]=38.01, p<.0001), education level (chi2[2]=11.5, p=.0032), and region (chi2[4]=66.1, p<.0001). These 3 variables were considered as covariates.

**Table 1.** Baseline characteristics of MASLIHAT trial participants (N=420)

|  | MASLIHAT |  | TANSIHAT |  |
| --- | --- | --- | --- | --- |
|  | Mean (SD) |  | Mean (SD) |  |
| Age | 29.7 (6.1) |  | 30.3 (6.3) |  |
| Monthly income (Rubles) <sup>a</sup> | 44294 (9501) |  | 42717 (9182) |  |
| **Education** | **N** | **%** | **N** | **%** |
| Primary | 9 | 4.3 | 7 | 3.3 |
| Secondary | 115 | 54.8 | 125 | 59.5 |
| College or Technical | 50 | 23.8 | 55 | 26.2 |
| University (but no degree) | 9 | 4.3 | 5 | 2.4 |
| University Degree | 27 | 12.9 | 18 | 8.6 |
| **Employment** |  |  |  |  |
| Construction | 114 | 54.5 | 116 | 55.5 |
| Loading in Bazaar | 41 | 19.6 | 46 | 22.0 |
| Selling in Bazaar | 34 | 16.3 | 22 | 10.5 |
| Food Service in Bazaar | 8 | 3.8 | 11 | 5.3 |
| Other | 12 | 5.7 | 14 | 6.7 |
| **Marital Status <sup>b</sup>** |  |  |  |  |
| Not Married | 100 | 47.8 | 76 | 36.4 |
| Married | 18 | 8.6 | 34 | 16.3 |
| Divorced | 91 | 43.5 | 99 | 47.4 |
| **Ever any STI** |  |  |  |  |
| No | 102 | 48.6 | 103 | 49.1 |
| Yes | 85 | 40.5 | 88 | 41.9 |
| Don't know | 20 | 9.5 | 18 | 8.6 |
| Not answered | 3 | 1.4 | 1 | 0.5 |
| **HIV Positive / Tested <sup>c</sup>** | **19 / 207** | **9.2** | **9 / 206** | **4.4** |

<sup>a</sup> $t = 2.42, p < .05$
<sup>b</sup> $\text{Chi}^2 = 8.53, p = .014$
<sup>c</sup> Tested after baseline survey
MASLIHAT: “Migrants’ Approached Self-Learning Intervention in HIV/AIDS for Tajiks” (intervention arm); TANSIHAT Targeted Application of Network and Social Intervention on Health Assistance for Tajiks (control arm); SD: standard deviation

Over 90% of participants completed all follow-up surveys. Thirty-seven participants (8.8%) were lost to follow-up at 9 (n=19) and 12 months (n=18). Loss to follow-up was similar across treatment arms and participant type. No new HIV infections were detected at 12-month follow-up. During the 12-month follow-up period 6.7% (n=14) of MASLIHAT participants and their network members and 19.0% (n=40) of TANSIHAT comparison arm participants and network members reported an STI diagnosis.

### Effects of intervention on sexual risk behaviors

Marginal contrasts of the arm by time interaction effects on sexual risk behaviors from mixed effects logistic and modified Poisson regression models are shown in Table 2. Complete results are available in an additional file (Additional File 2). The interaction effect was significant for all outcomes that were estimable. Site level clustering was significant and included only for sex with multiple partners. All outcomes decreased significantly in the MASLIHAT arm but not in the TANSIHAT arm, and decreases were sustained over 12 months.

**Table 2.** Marginal contrasts of treatment arm by time interaction on sexual risk behaviors during 12-month follow-up, mixed effects regressions^a^ (N=420, nobs=2043)

|  | Chi<sup>2</sup> [4 df] | p-value |
| --- | --- | --- |
| 1. Any condomless sex<sup>b</sup> | 67.32 | < .0001 |
| 2. Sex w/ multiple partners<sup>b</sup> | 24.60 | 0.0001 |
| 3. Any sex with sex workers<sup>b</sup> | 23.79 | 0.0001 |
| 4. Number of sex worker partners<sup>b</sup> | 17.15 | 0.0018 |
| 5. Condomless sex with sex workers<sup>c</sup> | -- | -- |
| 6. Condomless sex with casual or commercial partner<sup>c</sup> | 79.51 | < .0001 |

<sup>a</sup> Model 2 included random intercepts for participant, network cluster, and site. All other models included random intercepts for participant and network cluster. Covariates included participant type, marital status and income.
<sup>b</sup> modified Poisson regression model <sup>c</sup> logistic regression model
-- not estimable
MASLIHAT: “Migrants’ Approached Self-Learning Intervention in HIV/AIDS for Tajiks” (intervention arm); TANSIHAT Targeted Application of Network and Social Intervention on Health Assistance for Tajiks (control arm); CI: confidence interval

#### Effects of sexual risk behavior on reported STI

Results of the mixed effects logistic regression models testing the effects of treatment arm and sexual risk behavior on STI are shown in Table 3. Participant type, time, and time x arm effects were not significant predictors of STI and were dropped from the models. Region, marital status and income also were not significant predictors of STI during follow-up and were not included in subsequent models. Participants in the MASLIHAT condition were significantly less likely to report an STI during follow-up. The marginal probability of STI in the past 3 months over the 12-month follow-up period was 0.024 (95% CI 0.009 – 0.039) in the MASLIHAT condition and 0.086 (95% CI 0.055 – 0.116) in the TANSIHAT condition (conditional marginal effect dy/dx = -.062, p<.0001).

**Table 3.** Effects of treatment arm and sexual risk behaviors on STI during 12-month follow-up, mixed effects logistic regressions (N=420, nobs=1602)

|  | OR (95% CI) | p-value |
| --- | --- | --- |
| MASLIHAT vs. TANSIHAT | 0.14 (0.05, 0.39) | <0.001 |
| Education |  |  |
| College degree or some university | 1.38 (0.52, 3.63) | 0.519 |
| University degree | 4.82 (1.42, 16.39) | 0.012 |
| *vs. primary or secondary* |  |  |
| Employment sector |  |  |
| Loading in bazaar | 5.10 (1.70, 15.29) | 0.004 |
| Selling or food service in bazaar | 1.72 (0.42, 7.03) | 0.448 |
| Other occupation | 6.51 (1.46, 29.10) | 0.014 |
| *vs. construction* |  |  |
| Residual intraclass correlations | ICC | SE |
| Network cluster | .189 | .129 |
| Participant \| Cluster | .642 | .064 |
| **Sexual risk models†** |  |  |
| 1. Any condomless sex | 1.54 (0.59, 4.03) | 0.378 |
| MASLIHAT vs. TANSIHAT | 0.16 (0.06, 0.43) | <0.001 |
| 2. Sex w/ multiple partners | 1.73 (0.64, 4.70) | 0.284 |
| MASLIHAT vs. TANSIHAT | 0.16 (0.05, 0.45) | 0.001 |
| 3. Any sex with sex workers | 2.42 (0.98, 6.01) | 0.057 |
| MASLIHAT vs. TANSIHAT | 0.16 (0.05, 0.46) | 0.001 |
| 4. Number of sex worker partners | 1.45 (0.84, 2.49) | 0.178 |
| MASLIHAT vs. TANSIHAT | 0.16 (0.06, 0.48) | 0.001 |
| 5. Condomless sex with sex workers | 8.15 (2.30, 28.91) | 0.001 |
| MASLIHAT vs. TANSIHAT | 0.23 (0.08, 0.69) | 0.008 |
| 6. Condomless sex with casual or commercial partner | 4.02 (1.38, 11.72) | 0.011 |
| MASLIHAT vs. TANSIHAT | 0.25 (0.05, 1.30) | 0.098 |

† Each sexual risk behavior entered together with treatment arm and covariates education level and employment sector MASLIHAT: “Migrants’ Approached Self-Learning Intervention in HIV/AIDS for Tajiks” (intervention arm); TANSIHAT Targeted Application of Network and Social Intervention on Health Assistance for Tajiks (control arm); CI: confidence interval

Both CS/FSW and CS/CC were significantly associated with STI during follow-up, and the effect of treatment arm was reduced by 24 to 28% when each of these variables were added to the model. The prevalence of CS/FSW was extremely low in the MASLIHAT condition, with only one participant reporting this behavior at 3 or 6 months. Consequently, estimating regression models using CS/FSW as an outcome wasn’t possible and mediation analysis focused solely on CS/CC as an outcome.

### Causal Mediation Models

The results of the mediation model for CS/CC are reported in Table 4. Nineteen participants were lost to follow-up at 9 months leaving a sample size of n=401, with 137 network clusters. An additional two participants were missing information on covariates, leaving a sample size of n=399. Since attrition was equal across treatment arms and across 3-month CS/CC we did not use imputation. Effects of covariates should be interpreted with caution (not shown here, full results available in Additional File 2.) The interaction of baseline behavior with treatment arm was not statistically significant but was retained in the model so as not to overestimate the mediation effect. The indirect effect of treatment arm via CS/CC was statistically significant and accounted for an estimated 26% of the total effect.

**Table 4.**
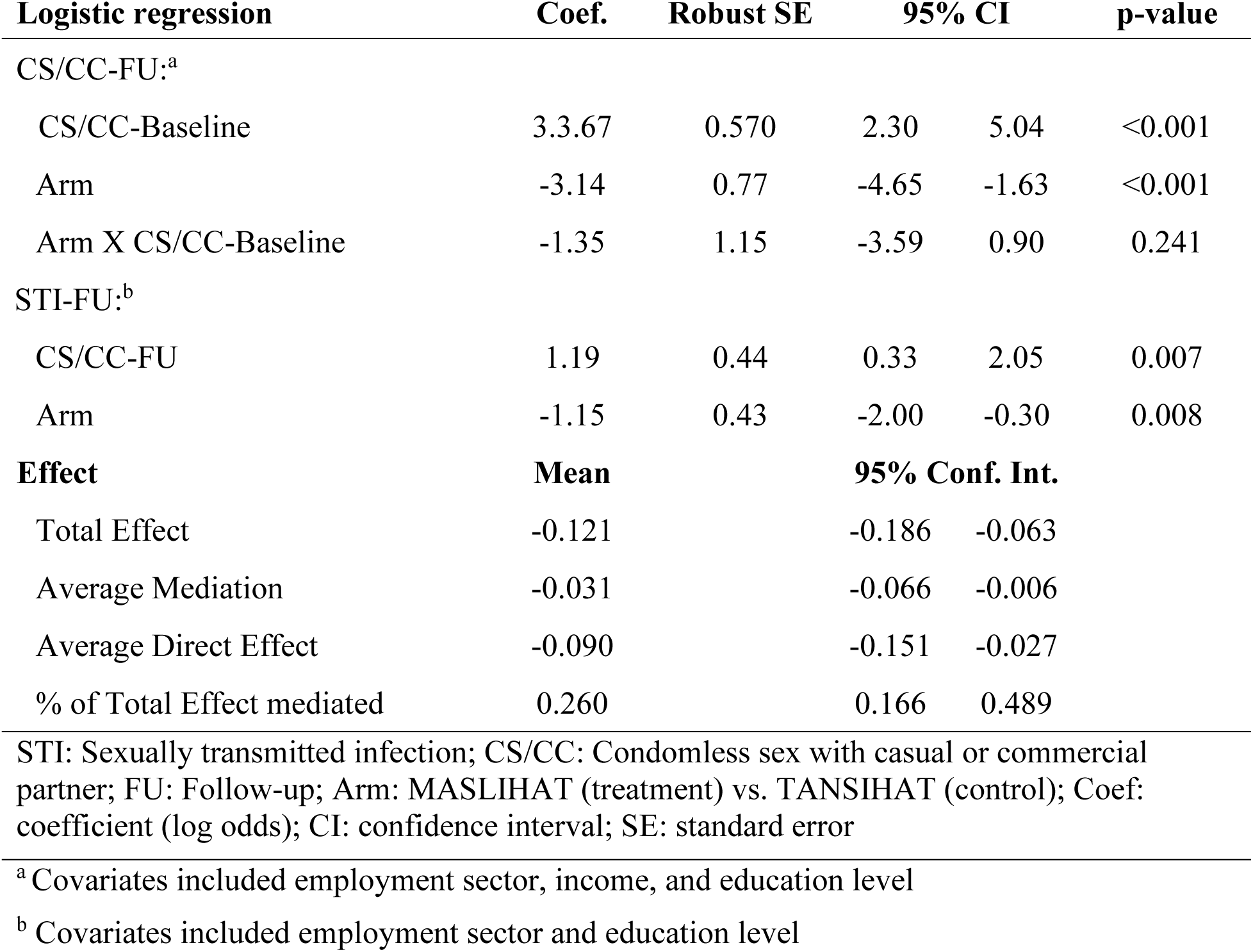
Causal mediation analysis testing effect of intervention arm on sexually transmitted infection via condomless sex with a casual or commercial partner (n=399)

## Discussion

The MASLIHAT intervention for HIV prevention is a network-based peer education intervention tailored for Tajik migrants who inject drugs while working in Russia. In previous analyses of this cluster-randomized controlled trial, we found significant reductions in self-reported sexual risk behaviors associated with the intervention including sexual activity with female sex workers and condomless sex. Consistent with our hypotheses, our findings indicate that STI diagnoses occurred less often during one year of follow-up among MASLIHAT PEs and network members compared to control arm PEs and network members. Moreover, the effect of the MASLIHAT intervention on STIs was partially mediated by changes in sexual activity, specifically engaging in condomless sex with casual and commercial female partners.

Risk for HIV and for STIs are behaviorally entwined as they share the same set of sexual practices that can lead to infection. While several indicators of sexual risk behavior were affected by the MASLIHAT intervention, our results highlight the importance of considering nuanced measures that capture behavior most likely to have an impact on sexual health. One-dimensional measures of condomless sex, commercial sex, and concurrent sexual partnering may have only weak associations with sexual health outcomes such as HIV and other STI acquisition. The most appropriate measures may also vary depending on the context. For male labor migrants, transactional sex - whether with sex workers or casual partners – is a common risk. (22)

Peer education interventions with people who inject drugs have demonstrated success in reducing risky injection behavior in a variety of populations,(19, 23–25) but effects on sexual behavior have not always been as clearly demonstrated.(23, 24, 26) The success of the MASLIHAT intervention in reducing risky sexual behavior may be tied to the cultural elements embedded in the peer education training that validate the migrant experience and help to increase the acceptability of having these discussions with their peers. If culturally adapted for use with other ethnicities, the intervention holds promise in reducing STIs among the many labor migrants in Moscow from other Central Asia countries and also PWID in other countries.

### Limitations

Our sample is composed entirely of male Tajik migrants who inject drugs while in Moscow and does not represent the sexual behavior of their Tajk counterparts who do not use drugs. Neither do the study’s findings represent the sexual behavior of the numerous labor migrants in Moscow from other Central Asian countries or the highly gendered experience of the increasing number of Tajik women in recent years who migrate to Russia for work.(27) The STI outcomes and sexual risk behaviors in this study are self-reported and cannot be verified. Consequently, it’s impossible to determine if participants’ responses have been influenced in any way by the demand characteristics of the intervention (28) or were subject to social desirability biases that inhibited disclosure of sensitive or embarrassing behavior.(29) Moreover most STDs are asymptomatic, making them difficult to detect without clinical confirmation or for individuals to recognize that they have contracted one.(30) Consequently, the rates of STI incidence reported by our study participants may under-report actual incidence of infection.

## Conclusions

The MASLIHAT peer-education intervention proved effective during a 12-month clinical trial in reducing STIs among Tajik labour migrant participants and their network members who inject drugs. As such, its findings contribute to WHO’s call for HIV/STI interventions and research findings that help to inform the goals of its widely adopted, “Global health sector strategies on HIV, viral hepatitis and sexually transmitted infections for the period 2022–2030.” MASLIHAT’s protective effects also may help to inhibit possible forward transmission of STIs among intervention participants to their wives and/or other sexual partners. If culturally adapted for other ethnicities, the intervention holds potential promise in reducing STIs among male migrant labour populations who inject drugs in other destination countries.

## Data Availability

The datasets supporting the findings of this study are available in the Open Science Framework repository. DOI 10.17605/OSF.IO/7G3YH.

https://osf.io/ws5mp/

## List of abbreviations

CS: condomless sex
CS/FSW: condomless sex with female sex worker
CS/CC: condomless sex with casual or commercial partner
FSW: female sex worker
HIV: Human Immunodeficiency Virus
MASLIHAT: Migrants’ Approached Self-Learning Intervention in HIV/AIDS for Tajiks
NM: Network member
PE: Peer educator
PWID: People who inject drugs
STI: Sexually transmitted infection
TANSIHAT: Targeted Application of Network and Social Intervention on Health Assistance for Tajiks
WHO: The World Health Organization

## Declarations

### Ethics approval and consent to participate

Study procedures were reviewed and approved by the Institutional Review Boards of the University of Illinois Chicago (protocol 2020-0795), PRISMA Research Center in Tajikistan, and the Moscow Nongovernment Organization “Bridge to the future.” All participants provided written informed consent.

### Consent for publication

Not applicable

### Availability of data and materials

The datasets supporting the conclusions of this article are available in the Open Science Framework repository [DOI 10.17605/OSF.IO/7G3YH https://osf.io/ws5mp/].

### Competing interests

The authors declare that they have no competing interests.

### Funding

This research was supported by a grant from the National Institute on Drug Abuse of the National Institutes of Health (USA) under Award Number R01DA050464 and by a grant from the National Center for Advancing Translational Science, NIH, through grant UL1TR002003. The content is solely the responsibility of the authors and does not necessarily represent the official views of the National Institutes of Health.

### Authors’ contributions

JAL and MEM contributed to the study’s conception and design. Material preparation and data collection were performed by JJ and CML. Data analysis was conducted by MEM and CML.

The first drafts of the manuscript were written by MEM and JAL and all authors commented on previous versions of the manuscript. All authors read and approved the final manuscript.

## Acknowledgements

We acknowledge the significant contributions of Dr. Mahbatsho Bahromov, founder of the PRISMA Research Center, that made this project possible. He passed away in February 2024. We thank the Tajik Diaspora Union, the Volunteer Doctors Association, and Moscow HIV Prevention Center for their assistance and the study’s participants and members of the MASLIHAT staff for making this research possible.

## Additional material

Additional File 1.pdf: Figure S1: CONSORT Diagram: Cluster Randomized Trial

## Notes

### Competing Interest Statement

The authors have declared no competing interest.

### Clinical Trial

NCT04853394

### Author Declarations

The University of Illinois Chicago Institutional Review Board, the PRISMA Research Center Institutional Review Board and the Moscow NGO "Bridge to the future" Institutional Board gave ethical approval for this work

### Summary of Updates

Substantive revisions were made based on reviewer feedback. The conclusions were unchanged.

