## Supplementary figures and images for "Peer Education Intervention Reduced Sexually Transmitted Infections Among Male Tajik Labor Migrants Who Inject Drugs: Results of a Cluster-randomized Controlled Trial"

### Additional File 1

**FIGURE S1. CONSORT DIAGRAM: CLUSTER RANDOMIZED TRIAL**

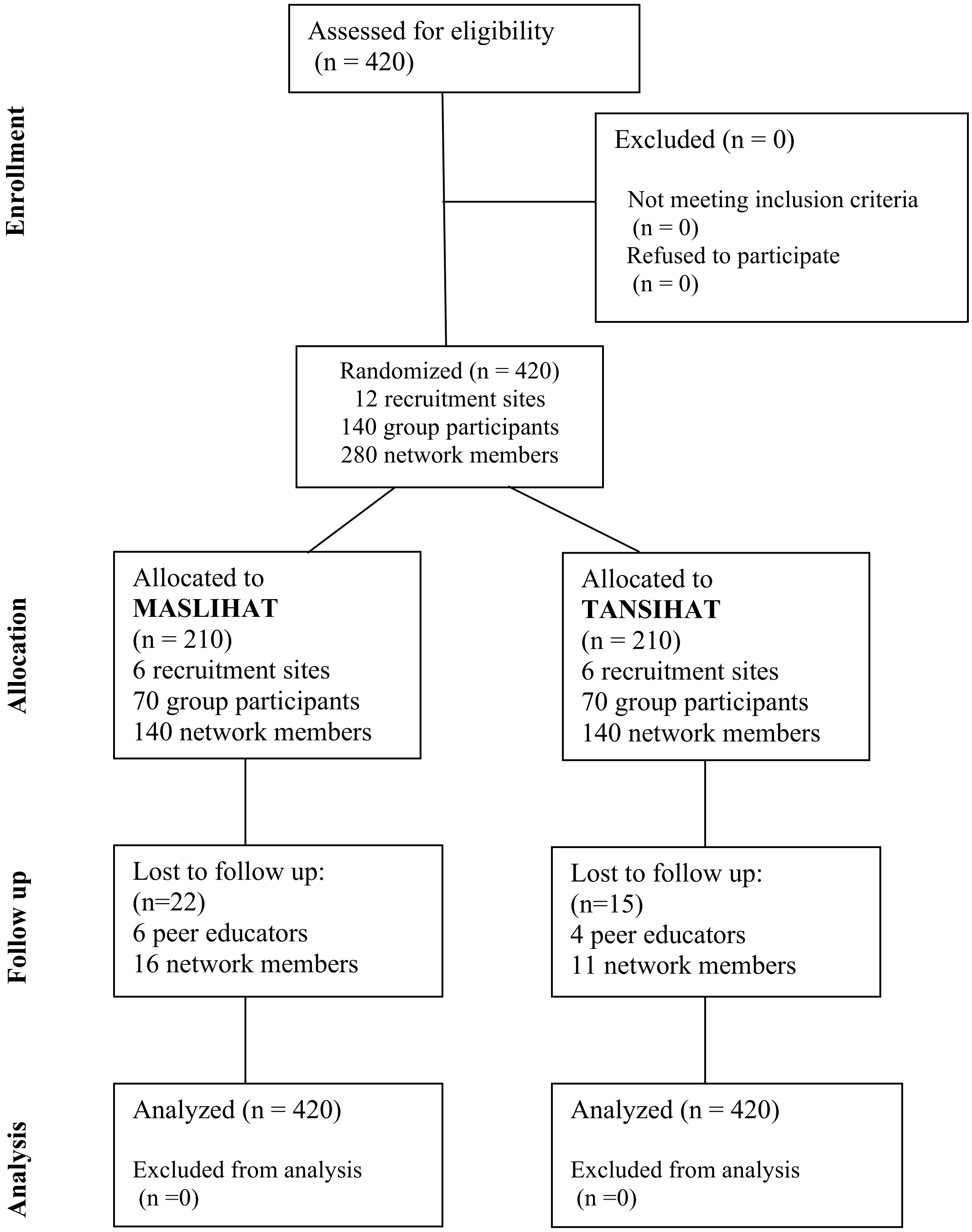
